# Primary Care for Homeless Veterans: A Systematic Review of the Homeless Patient Aligned Care Team (HPACT)

**DOI:** 10.1101/2021.03.30.21254619

**Authors:** David Rosenthal, Benjamin A. Howell, Erin Johnson, Katherine Stemmer Frumento, Jack Tsai

**Affiliations:** Yale School of Medicine, Department of General Internal Medicine, New Haven CT; 4catalyzer Inc., Guilford CT; Yale School of Medicine, National Clinician Scholars Program, New Haven, CT; VA National Center on Homelessness Among Veterans, Veterans Health Administration, Office of Homeless Programs, Washington D.C.; Yale School of Medicine, Harvey Cushing/John Hay Whitney Medical Library, New Haven, CT; University of Texas Health Science Center at Houston, School of Public Health, San Antonio, TX

**Author notes:** Corresponding author: David Rosenthal, M.D., Yale School of Medicine, 333 Cedar St, New Haven CT 06510.

**Keywords:** homelessness, primary care, Veterans

## Abstract

**Background:** In 2011, the Veterans Health Administration (VHA) implemented homeless-tailored primary care medical home models, called the Homeless Patient Aligned Care Teams (HPACT) to improve care for homeless Veterans. The aim of this study was to describe the existing peer-reviewed literature on HPACTs by systematically reviewing studies published since 2011 to date.

**Methods:** We conducted a systematic review of peer-reviewed studies published from 2011 to June 2019 to evaluate the literature since the inception of the VHA’s Homeless PACT program implementation. We included original research articles evaluating the Homeless PACT and excluded those that did not contain original data.

**Results:** Of 379 studies screened, 20 studies met our inclusion criteria and were included for analysis. Given wide variability in research designs and outcome measures, a narrative review was conducted. The 20 included studies were categorized into 3 groups: Early HPACT pilot implementations; Association of HPACT clinics with quality and utilization; and Specialized programs within HPACTs. Observational findings suggest reductions in emergency department utilization, improvements in primary care treatment utilization, engagement, and patient experience; but limited rigorous studies exist beyond single site pilots and a few large observational cohort studies.

**Discussion:** The HPACT model has been successfully implemented in VHA medical centers throughout the country with multiple studies showing increased primary care engagement and improved patient experience; however, further studies are needed about quality, utilization and whether the model can be implemented outside the VHA system.

**Registration Number/Funding source:** none

## Introduction

Homelessness is associated with premature morbidity and mortality at much higher rates compared to those who are housed. (1, 2) Health care for high-need, socially complex patients such as those persons experiencing homelessness is often inadequate, with high rates of acute healthcare utilization in the emergency department (3) decreased utilization of primary care, (4) and poor quality outcomes. (5, 6) The care provided to persons experiencing homelessness is often fragmented and reactive to acute presentations. Primary care, with its longitudinal, comprehensive, relationship-based model, in theory provides a platform for improved care and outcomes among those experiencing homelessness. Primary care has been described as “integrated, accessible health care services by clinicians who are accountable for addressing a large majority of personal health care needs, and practicing in the context of family and community.” (7) However, traditionally structured primary care models often do not meet the needs of socially vulnerable patients experiencing homelessness due to issues related to 1) trust, 2) stigma, and 3) care processes. (8)

In 2009, Secretary of Veterans Affairs (VA) Eric Shinseki unveiled the department’s comprehensive plan to end homelessness among Veterans. (9) Numerous Veterans Health Administration (VHA) programs were developed or expanded in response to this federal initiative. One of those programs was a population-tailored patient-centered medical home (known in VHA as a Patient-Aligned Care Team) for Veterans experiencing homelessness called the Homeless Patient Aligned Care Team (HPACT), which was implemented in select VA medical centers in 2011. Building on existing VHA primary care services, the intent of the HPACT model is to adapt the standard care delivery model to further integrate and coordinate health and social services care for homeless Veterans, focusing on the highest-risk, highest-need Veterans unable or unwilling to access traditional health care. (10) The HPACT model provides low-barrier, multidisciplinary care featuring open-access, wrap-around services and supports. As designed, the HPACT model distinguishes itself from traditional primary care settings by offering five core elements: 1) enhanced access, 2) integrated services addressing social determinants of health, 3) intensive care management to coordinated with community agencies, 4) specific training for providers on homeless care skills, and 5) data-driven accountable care processes using specific data reports to inform team performance on quality and utilization. Enhanced care entails adopting both an open-access, walk-in care model, reducing traditional barriers to receiving care, and outreach into the community to engage veterans disconnected with VA services. Service integration allows for one-stop, wrap around services to connect patients to mental health and homeless services, which are often co-located, given the often multiple social needs of veterans experiencing homelessness. In addition, primary care staff are able to provide food, clothing assistance, hygiene items, shower and laundry facilities which all serve to meet the continuum of veteran needs.(11)

The program was formally launched in 2011, starting with 32 sites and subsequently expanded to over 60 medical facilities and caring for approximately over 18,000 veterans annually nationwide, and many others who have since “graduated” to traditional VA primary care team—namely for those Veterans who have become stably housed and were able to navigate traditional VA primary care settings. Demographic data from the first 14,088 patients enrolled in HPACTs shows that the average participant was 53.4 years of age; 8.8% served in the military after September 11, 2001, 4.0% were women, and 11.0% were 65 years or older. (12) In parallel with the implementation and growth of the HPACT model, there have been many studies investigating various aspects of the HPACT model; however, no systematic review of this research in this area has yet been conducted. There is much to be learned from a review of published evaluations of the HPACT program. Specifically, a review would provide a summary and broader understanding about what is known about the HPACT Program, and to identify gaps in our understanding about the program. It may also provide insights and lessons learned for health systems caring for vulnerable populations outside the VA. This is a systematic review of the peer-reviewed literature to date.

## Methods

### Literature Search

This systematic review was conducted according to Preferred Reporting Items for Systematic Reviews and Meta-Analyses (PRISMA) guidelines. (13) We conducted a search using Medline, Embase, PsycINFO, Scopus, Web of Science, and Cochrane databases using the search strategy outlined in the accompanying appendix. Our search was limited to studies published in peer-reviewed literature.

### Study Selection

Articles were screened by two reviewers (D.R, B.H.) and included if they (1) evaluated an HPACT, (2) contained original data, and (3) were published after the implementation start of the HPACT program in 2011. As per figure 1 in the accompanying appendix, 379 studies were imported for screening, after removal of duplications, 371 studies were screened by two reviewers (D.R, B.H.) with 20 full text studies included for analysis. Each full-text article was reviewed by a pair of investigators (D.R, B.H). Disagreements were resolved by consensus.

### Data Extraction and Narrative Synthesis

For studies that met our inclusion criteria we extracted information on study objective, methodology, intervention, population, and relevant findings. Given the small number of studies and heterogenous methods, we conducted a narrative synthesis to describe our findings. Each article was reviewed in terms of research design, methods, and results. In attempt to pull out common themes across studies, we grouped studies into categories based on a combination of research design and results. These categories were determined by consensus of team members after reviewing the content of the included studies.

## Results

Ultimately, 20 studies were included in the review. Given the small number of studies, and the variability in research design and outcomes, we conducted a narrative systematic review of the included studies. We categorized the 20 included studies into three groups based on research design and content area. These groups were: early HPACT pilot implementations (five studies), association of HPACT Clinics with quality and utilization (nine studies), and specialized programs within HPACTs (six studies).

**See Table 1 (Attached)**

**Table 1.**
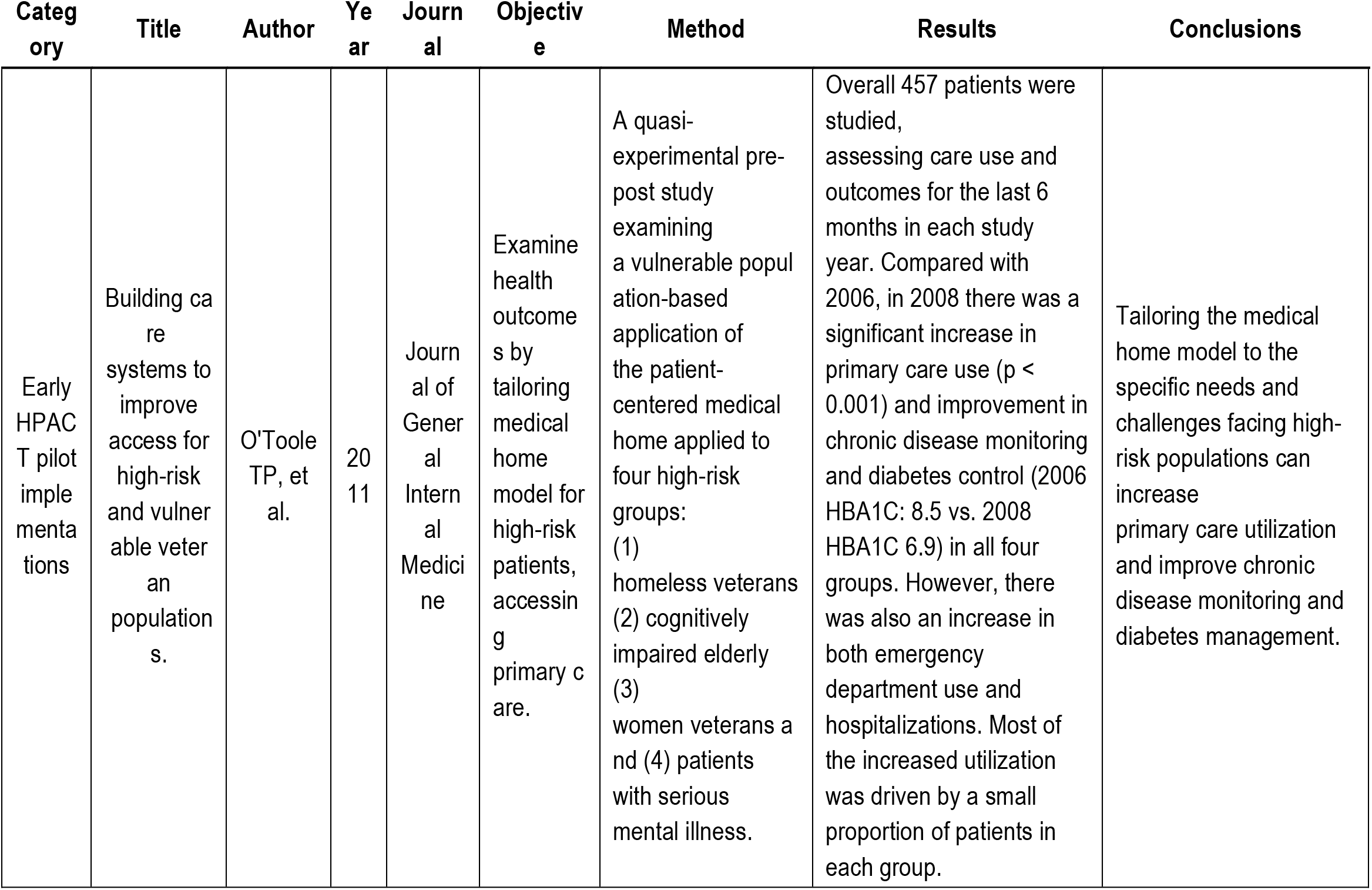
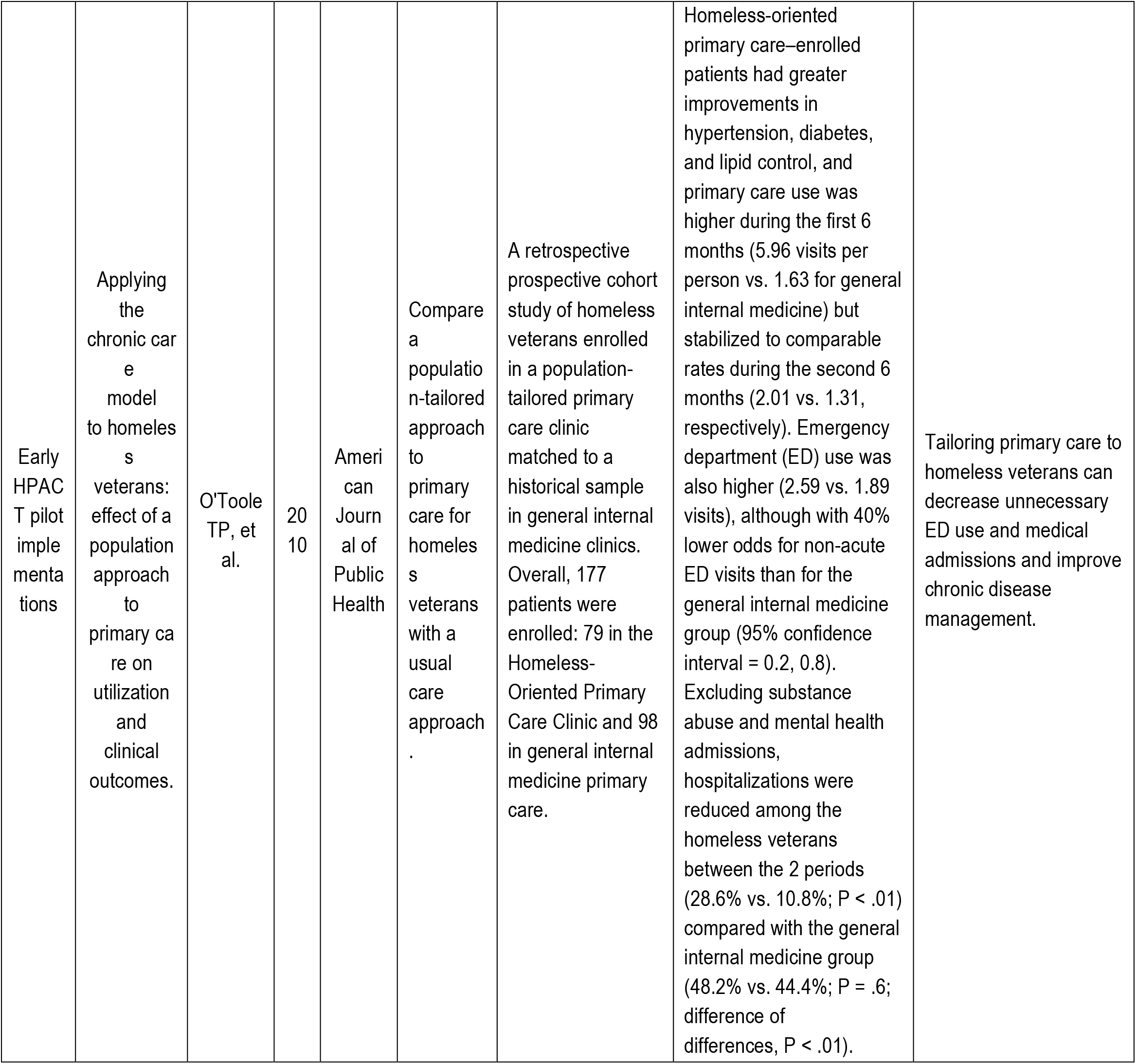
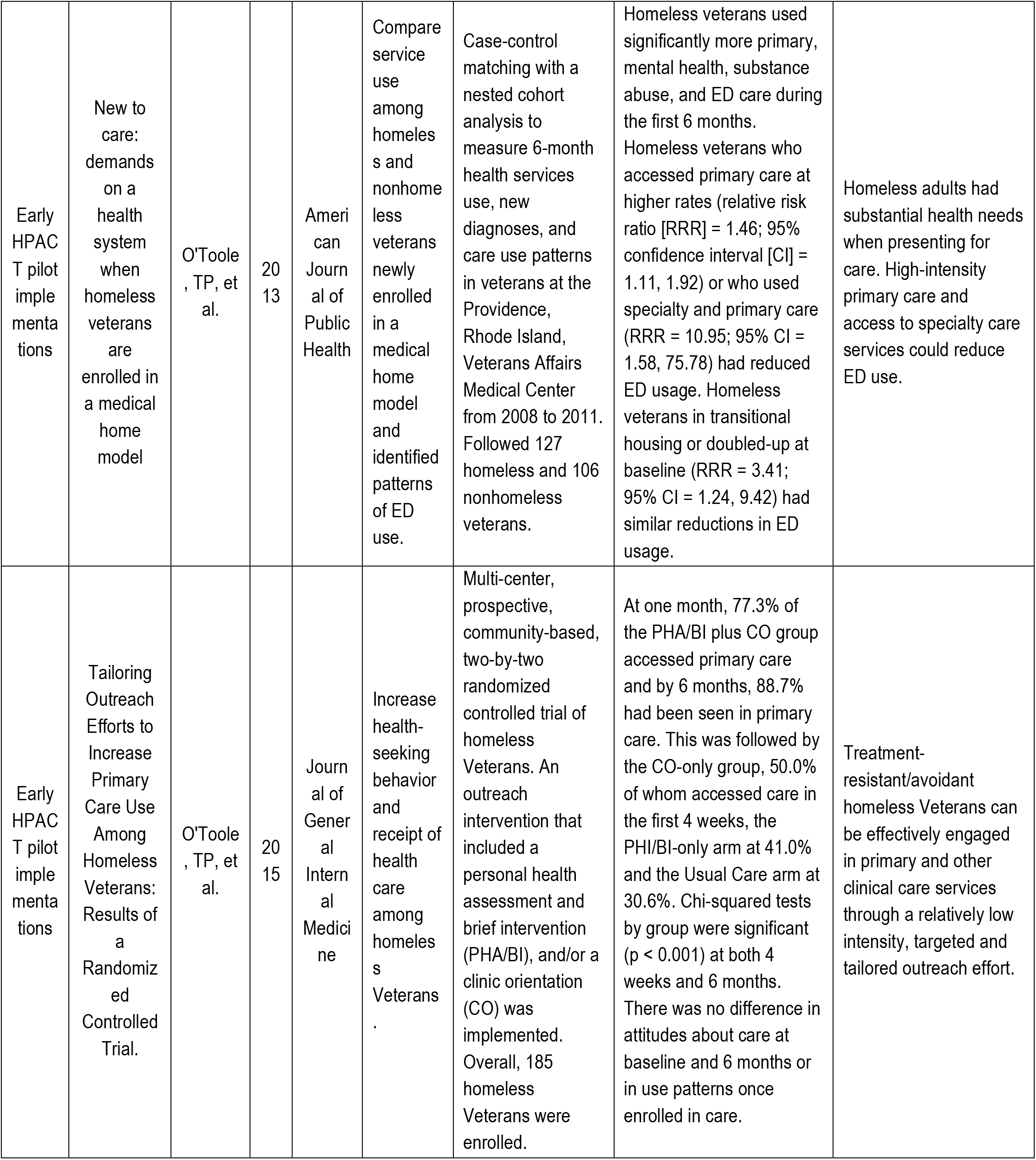
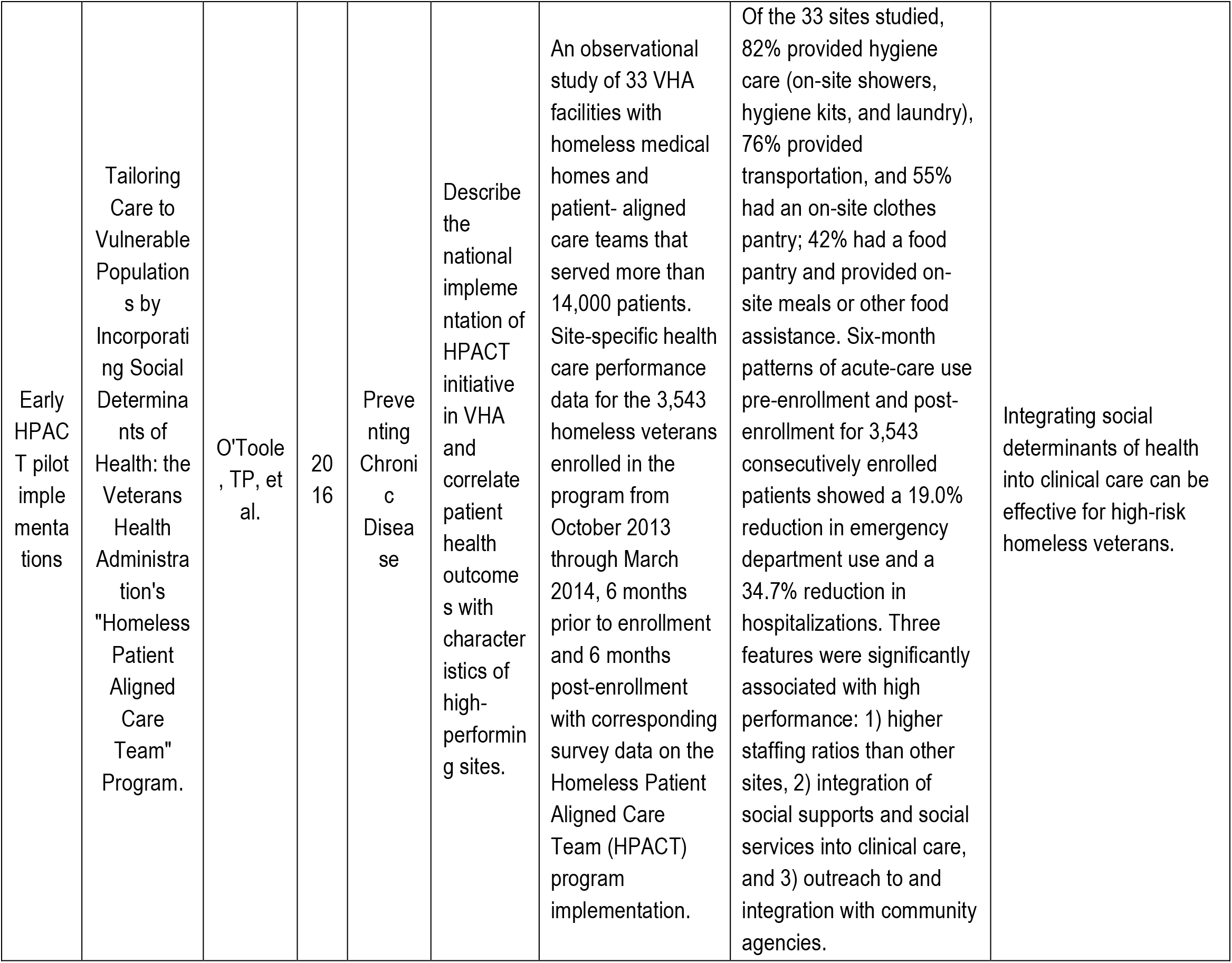
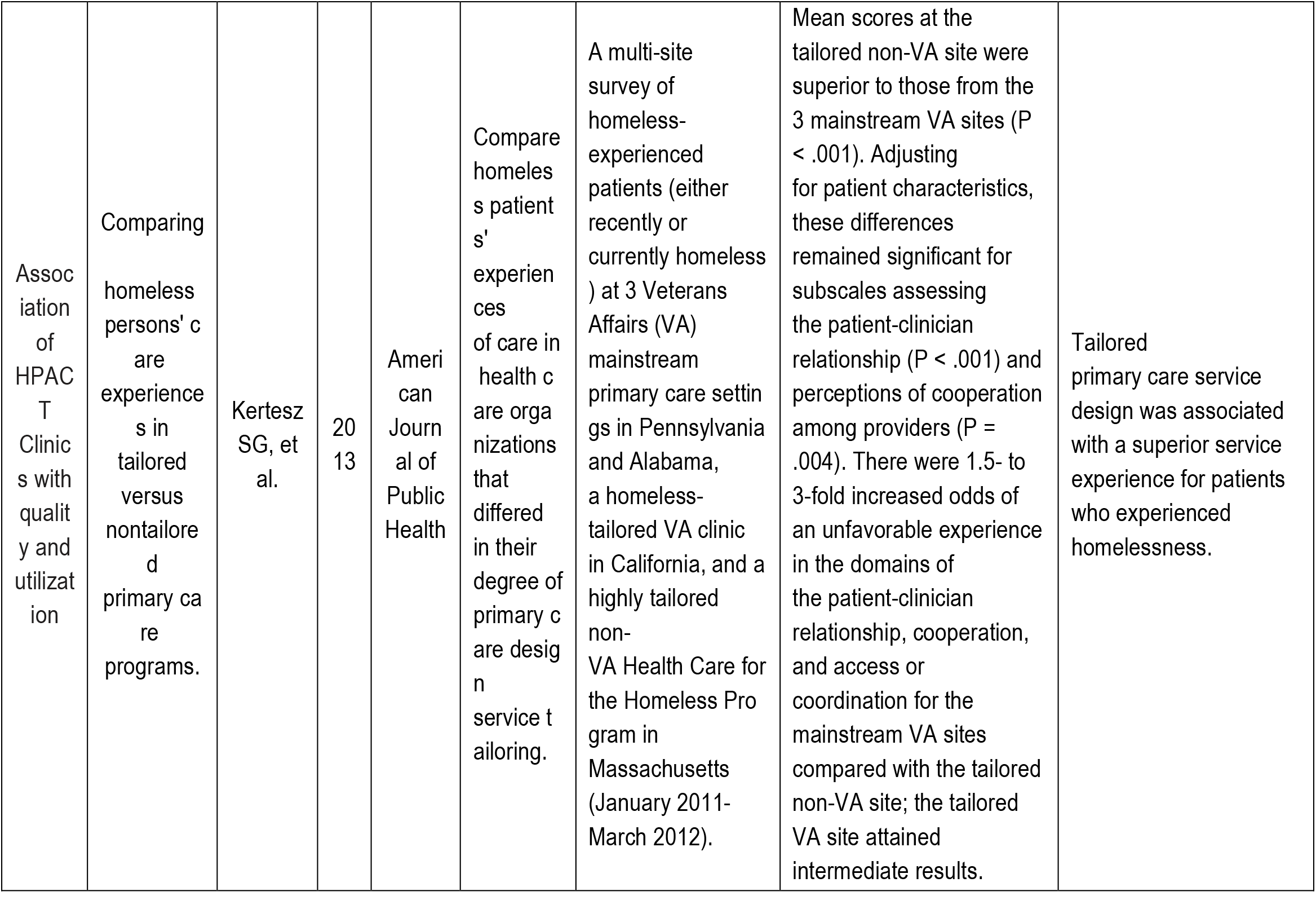
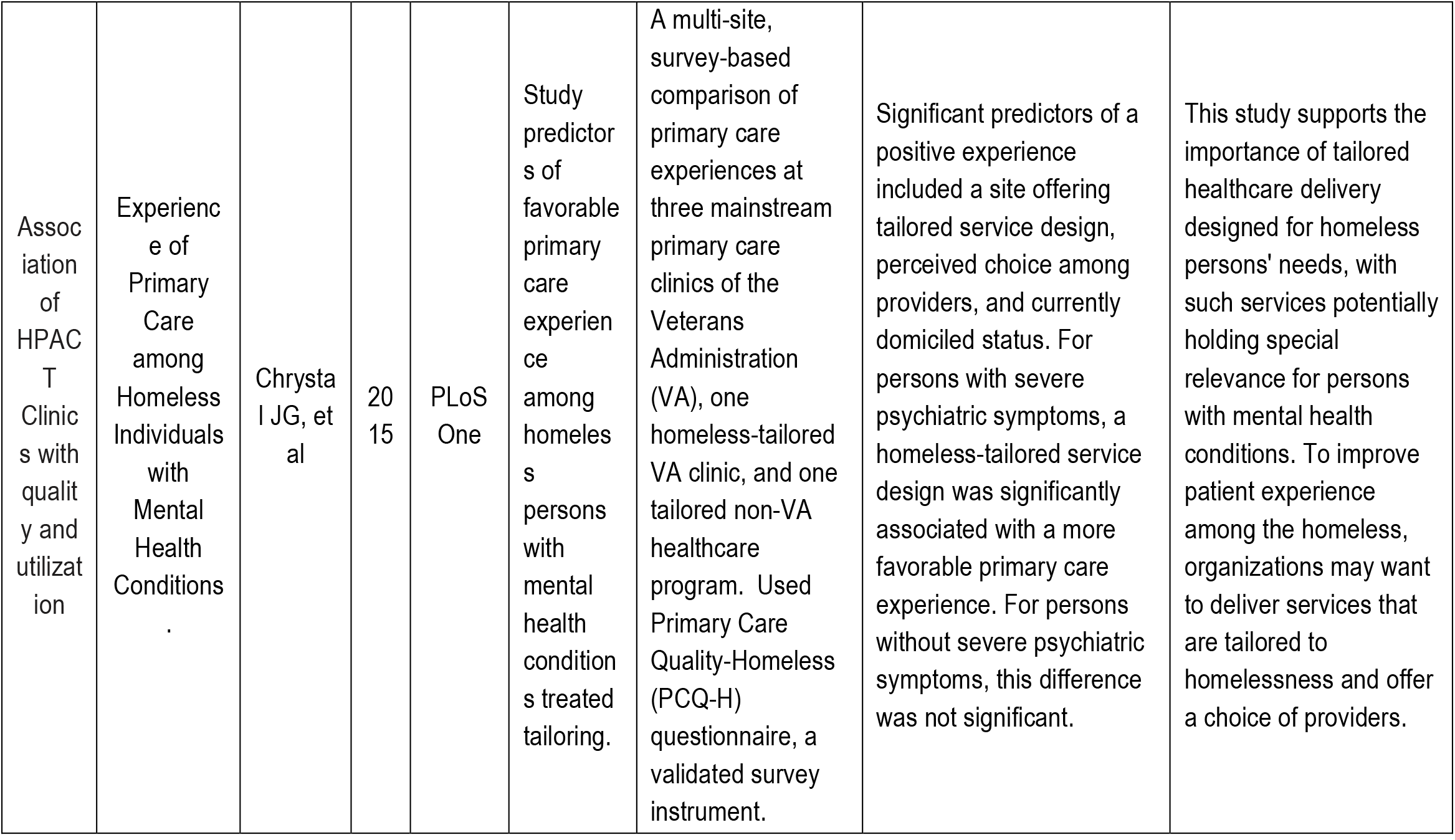
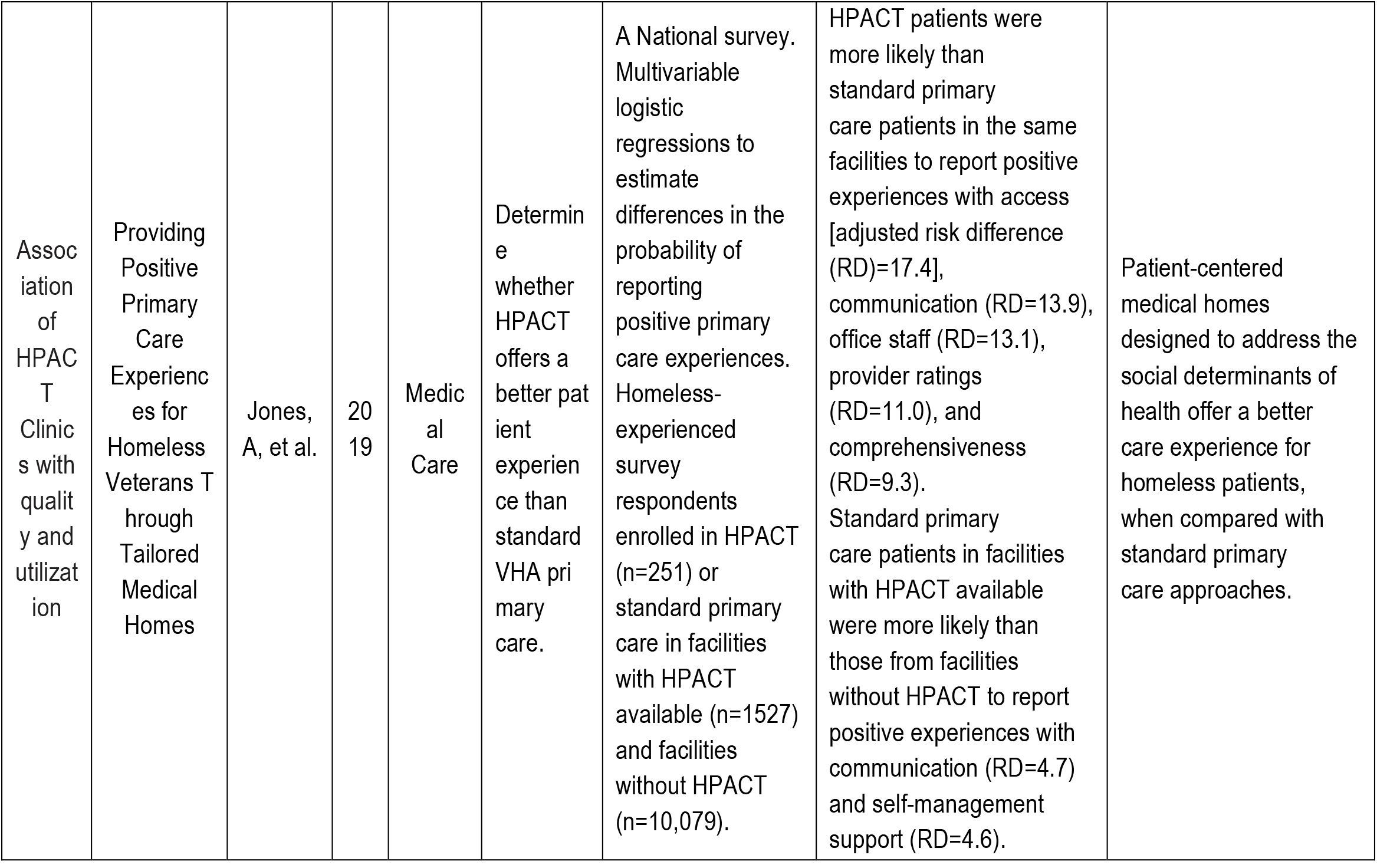
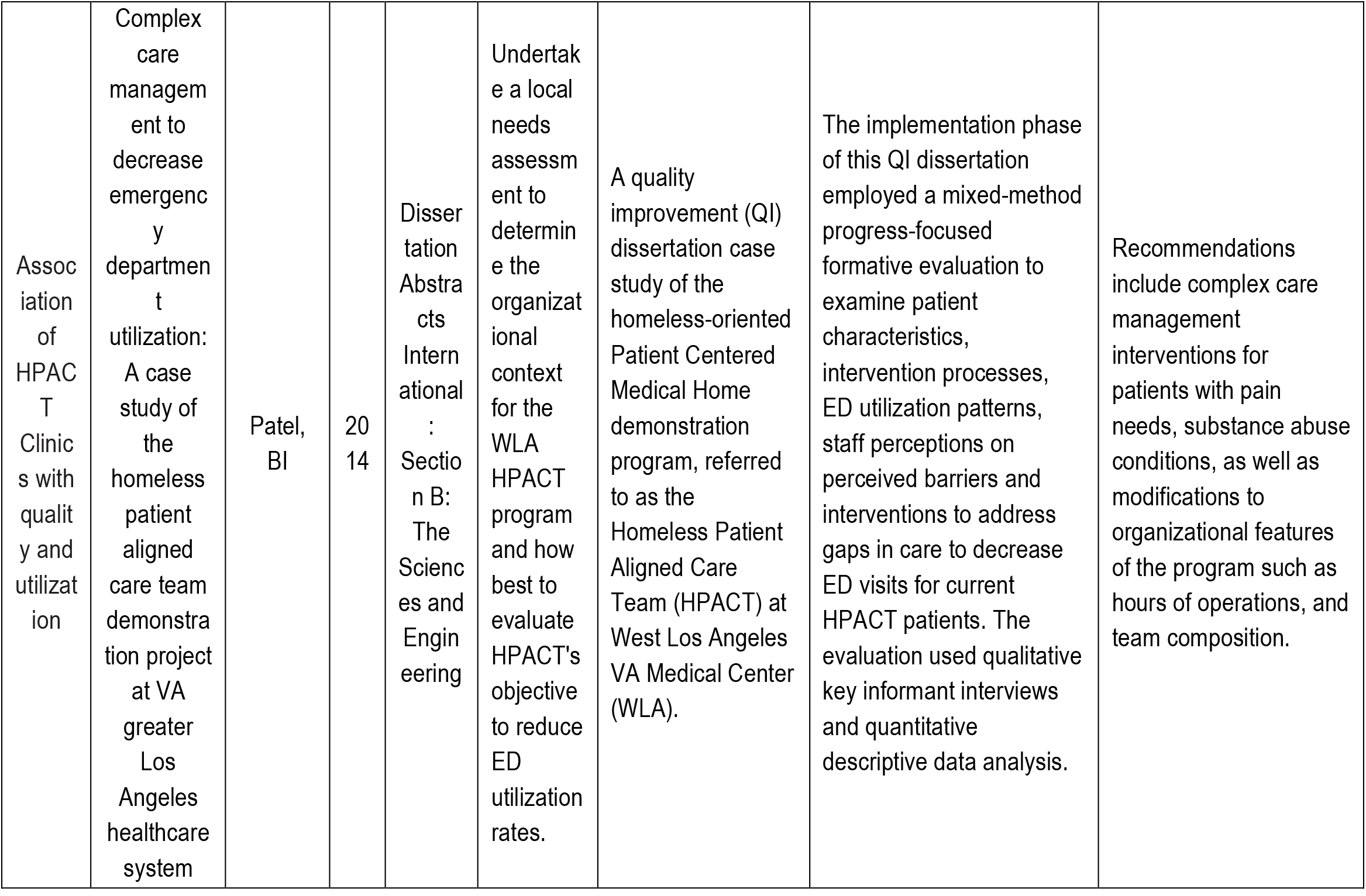
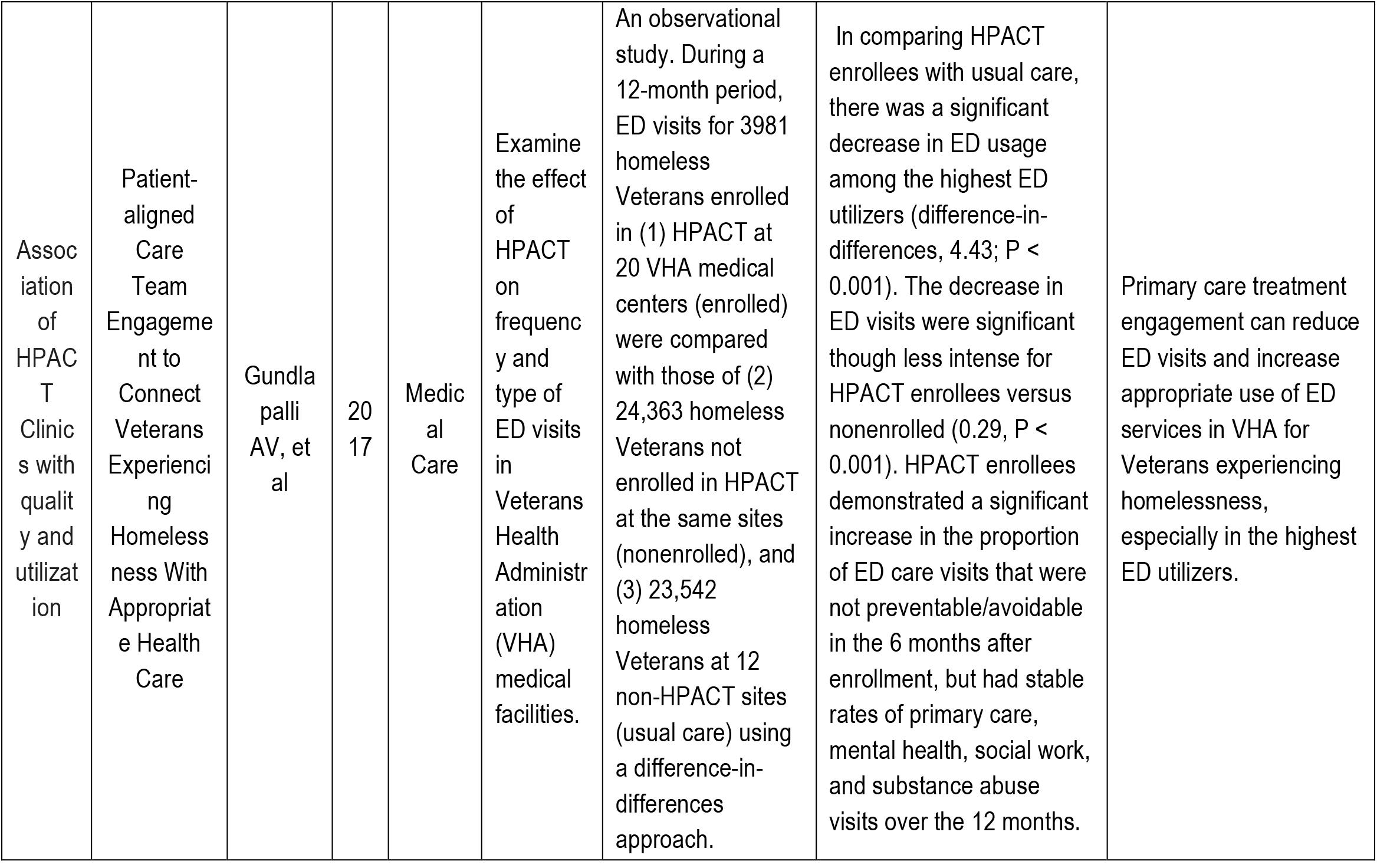
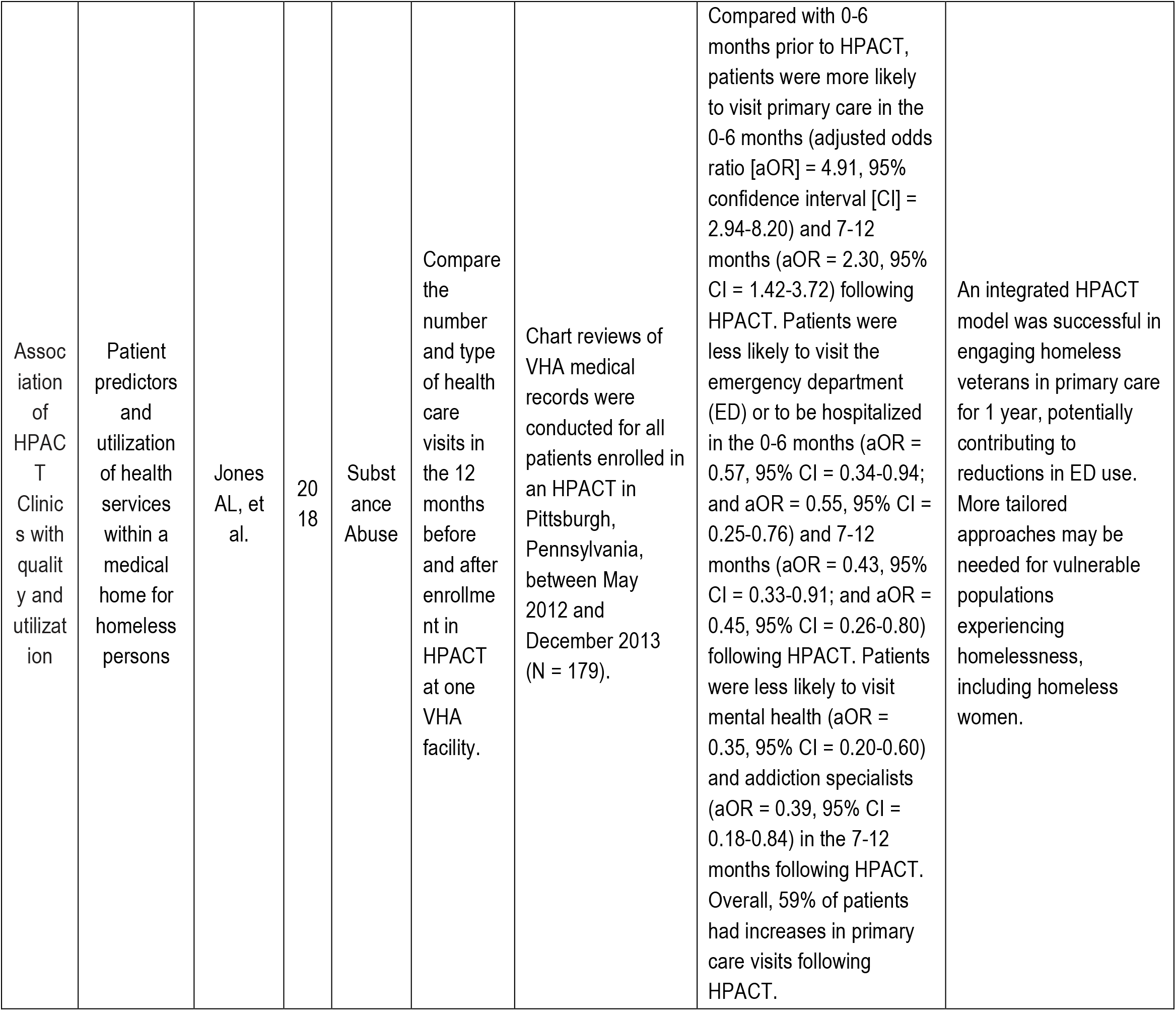
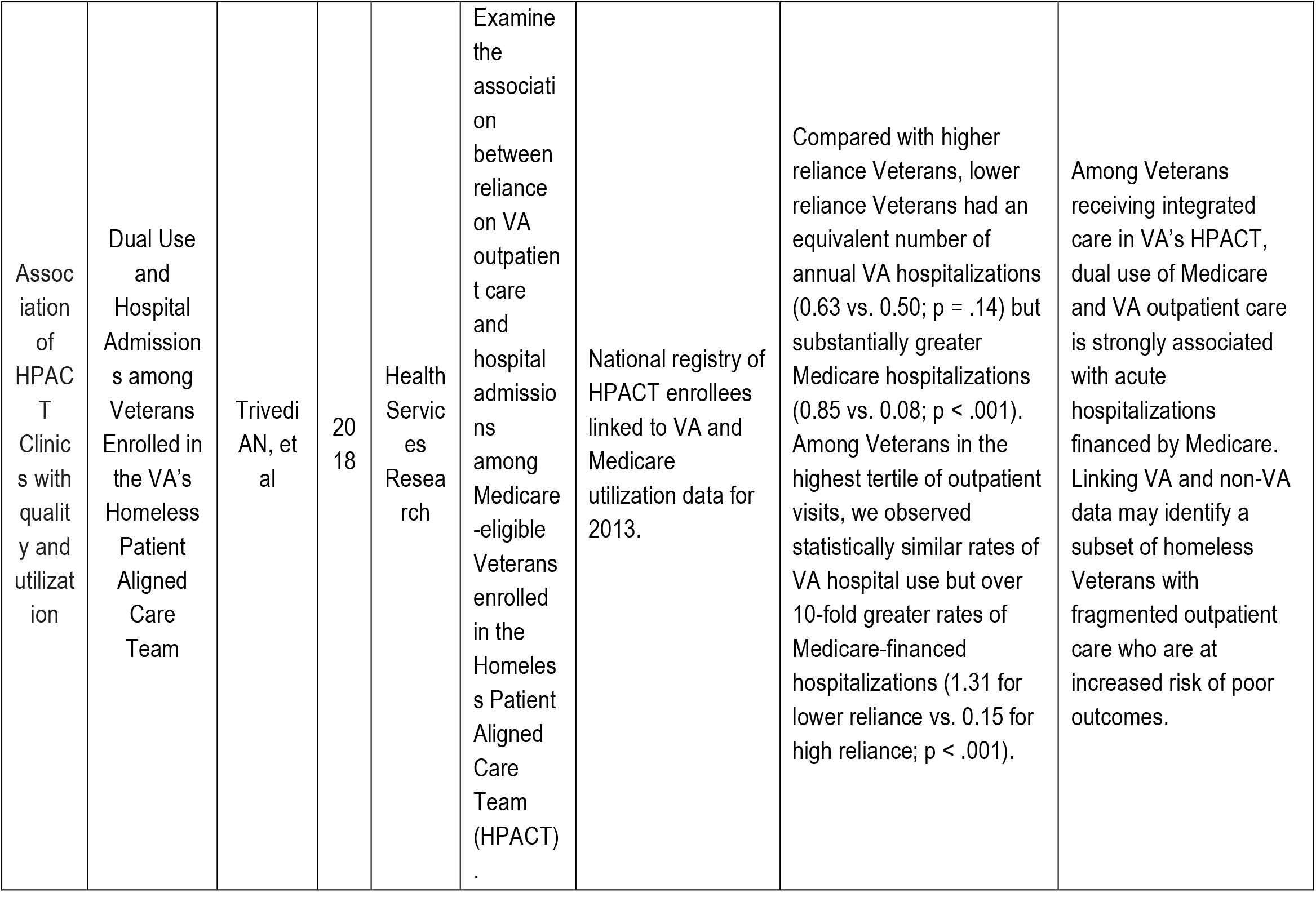
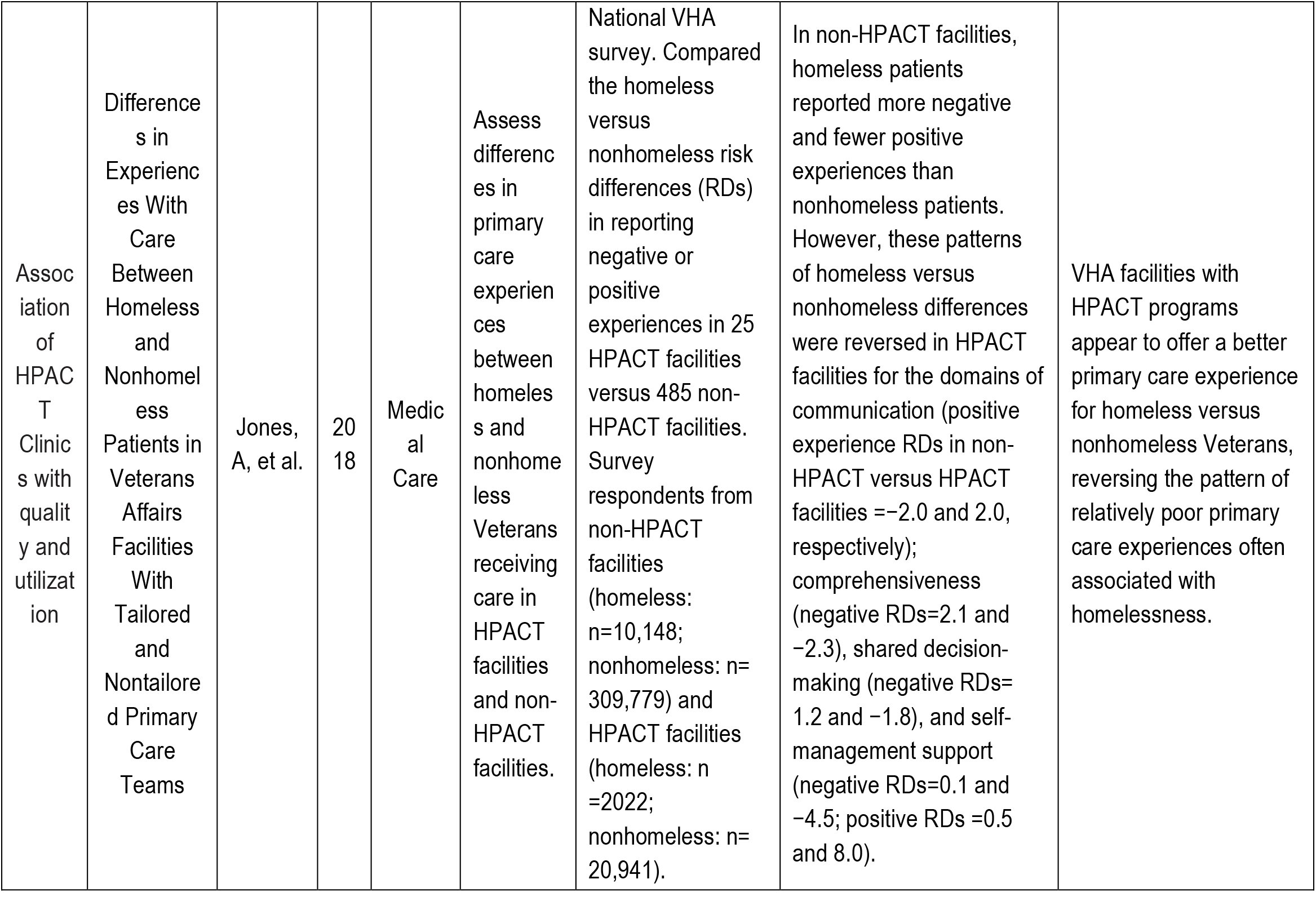
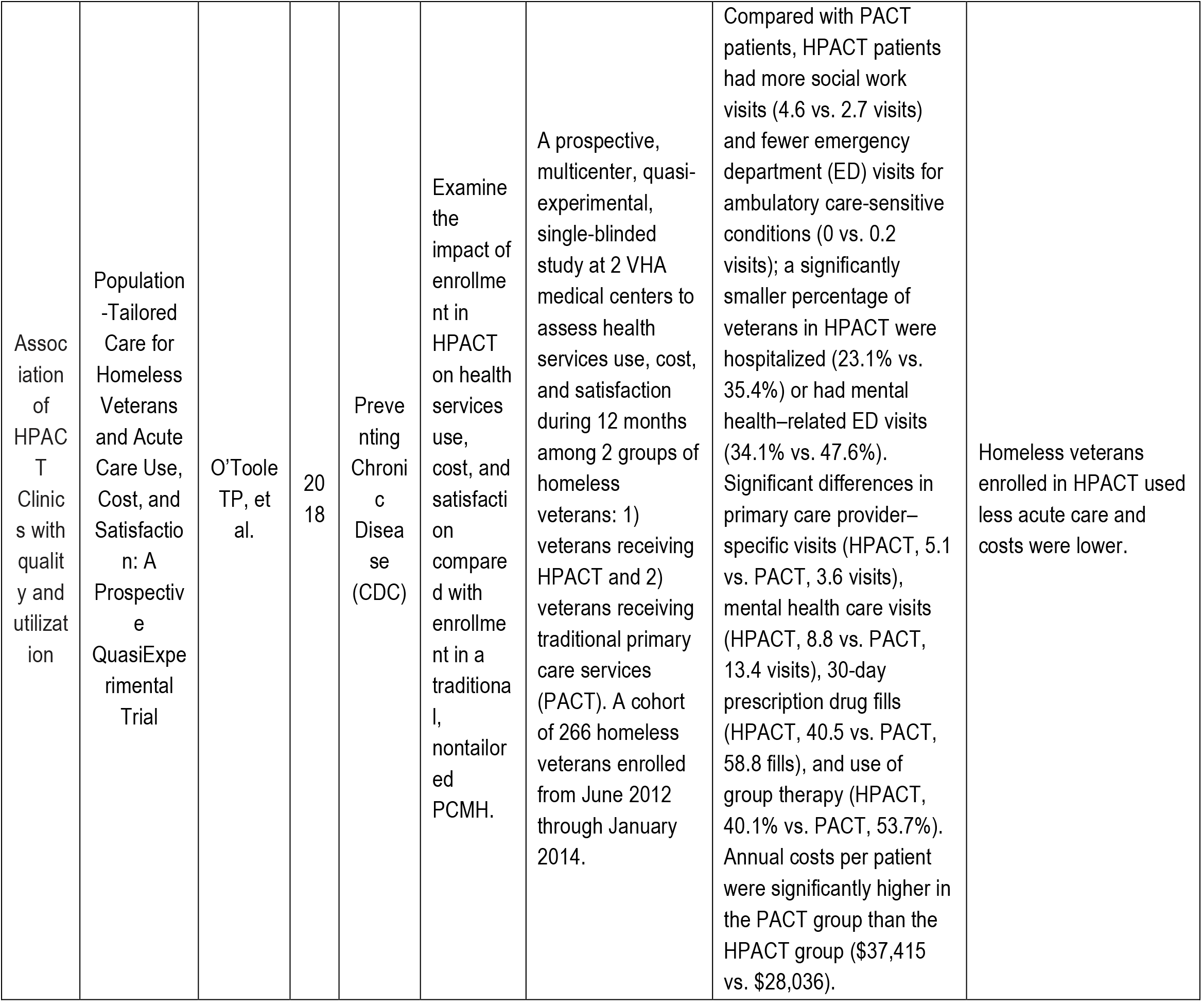
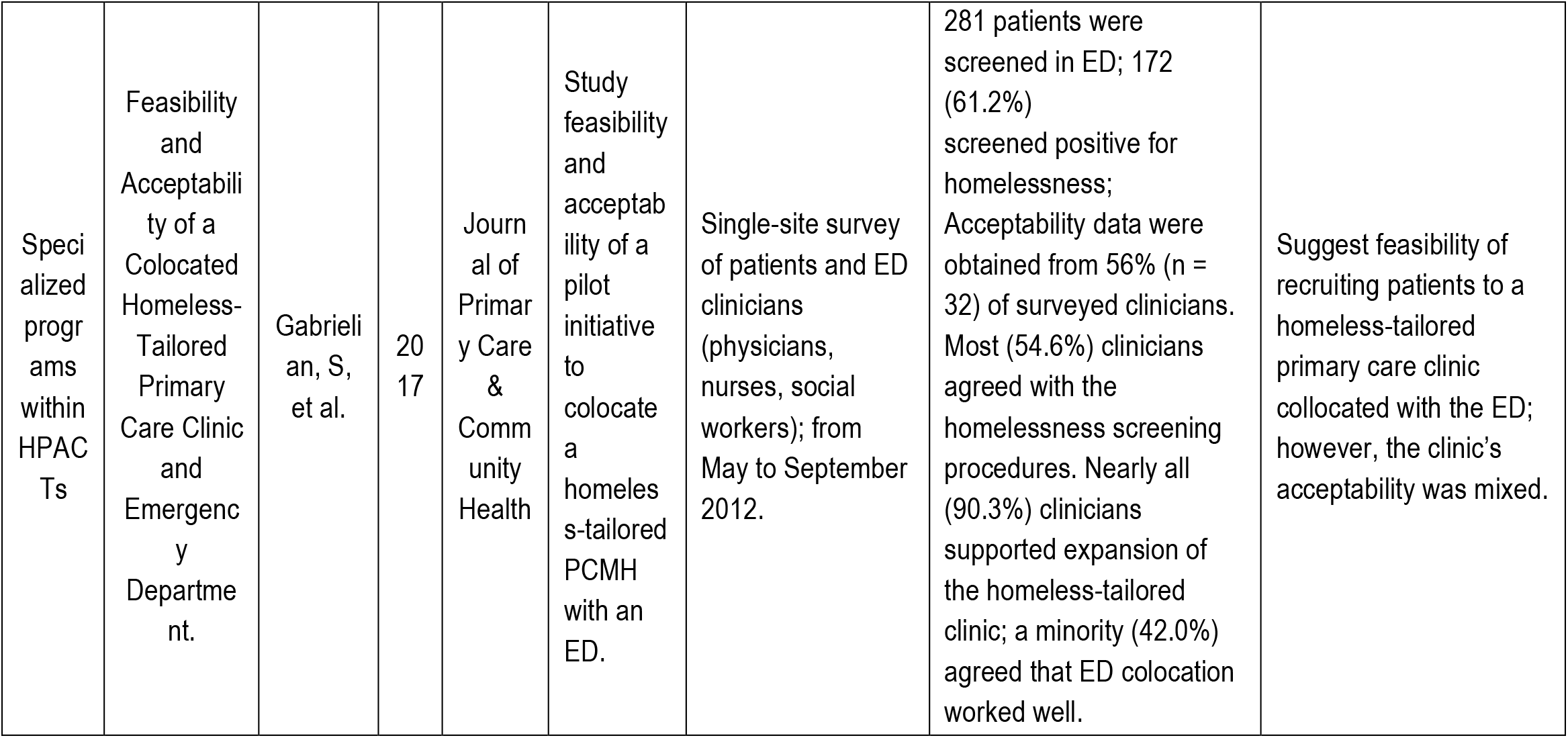
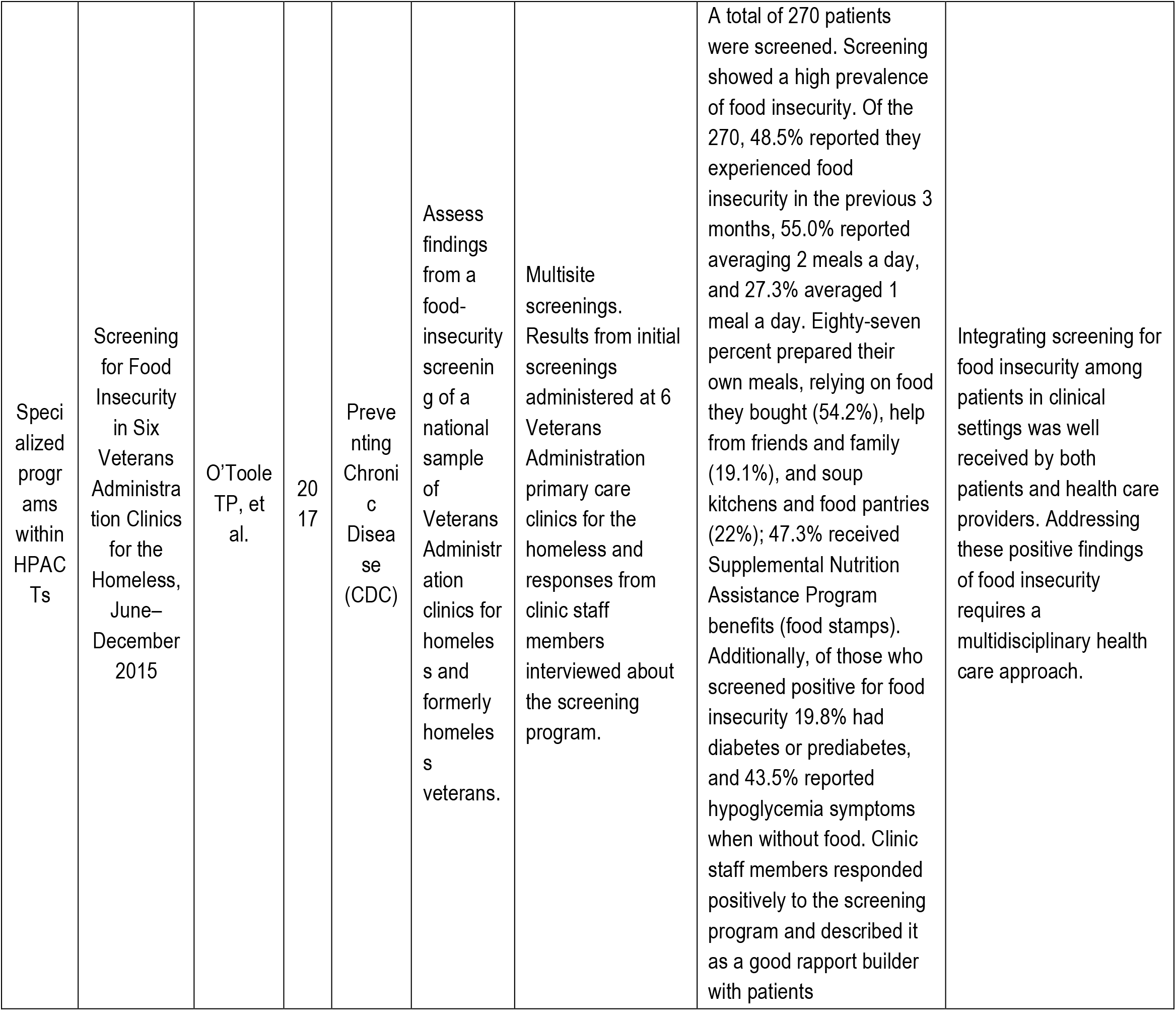
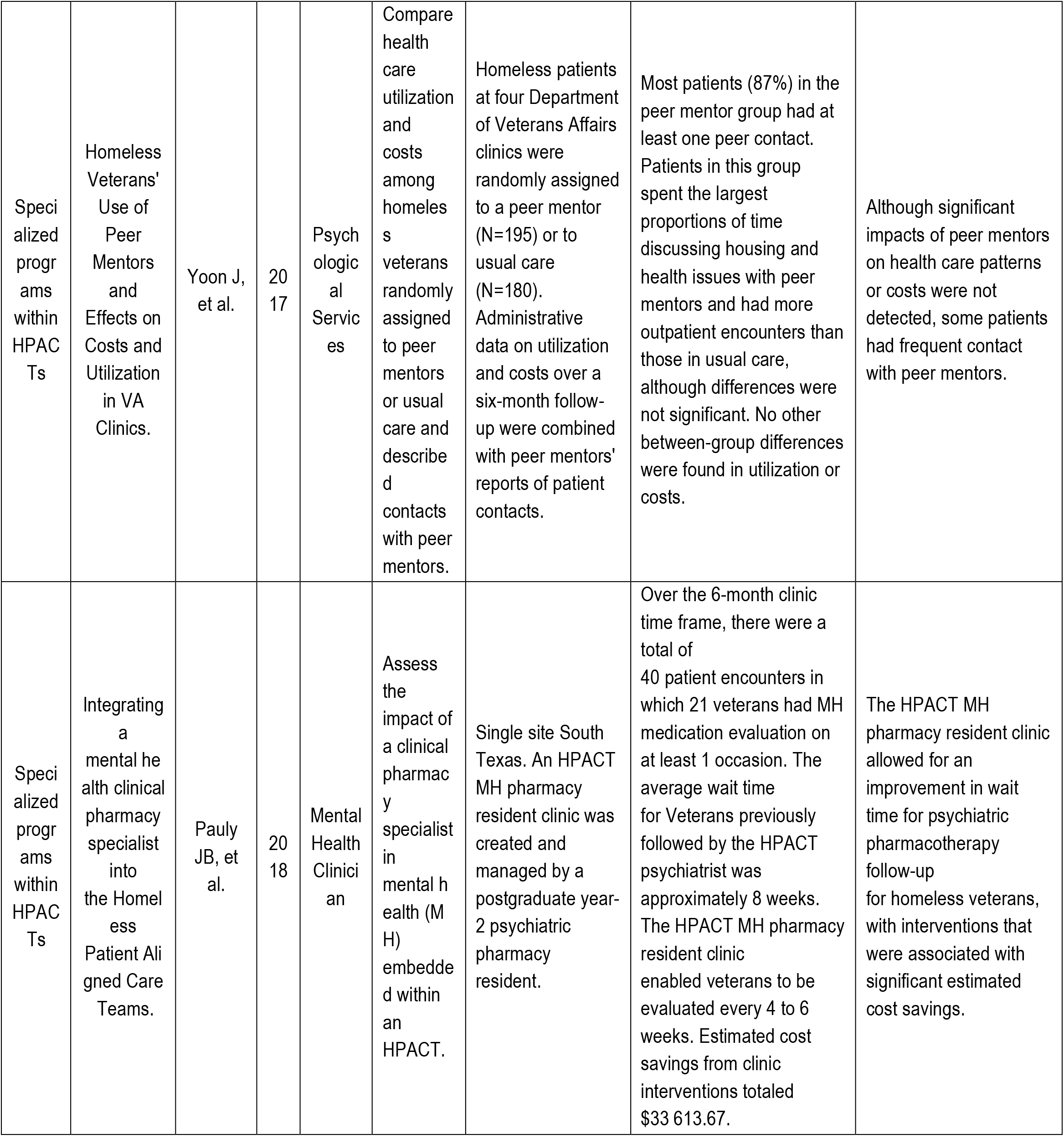
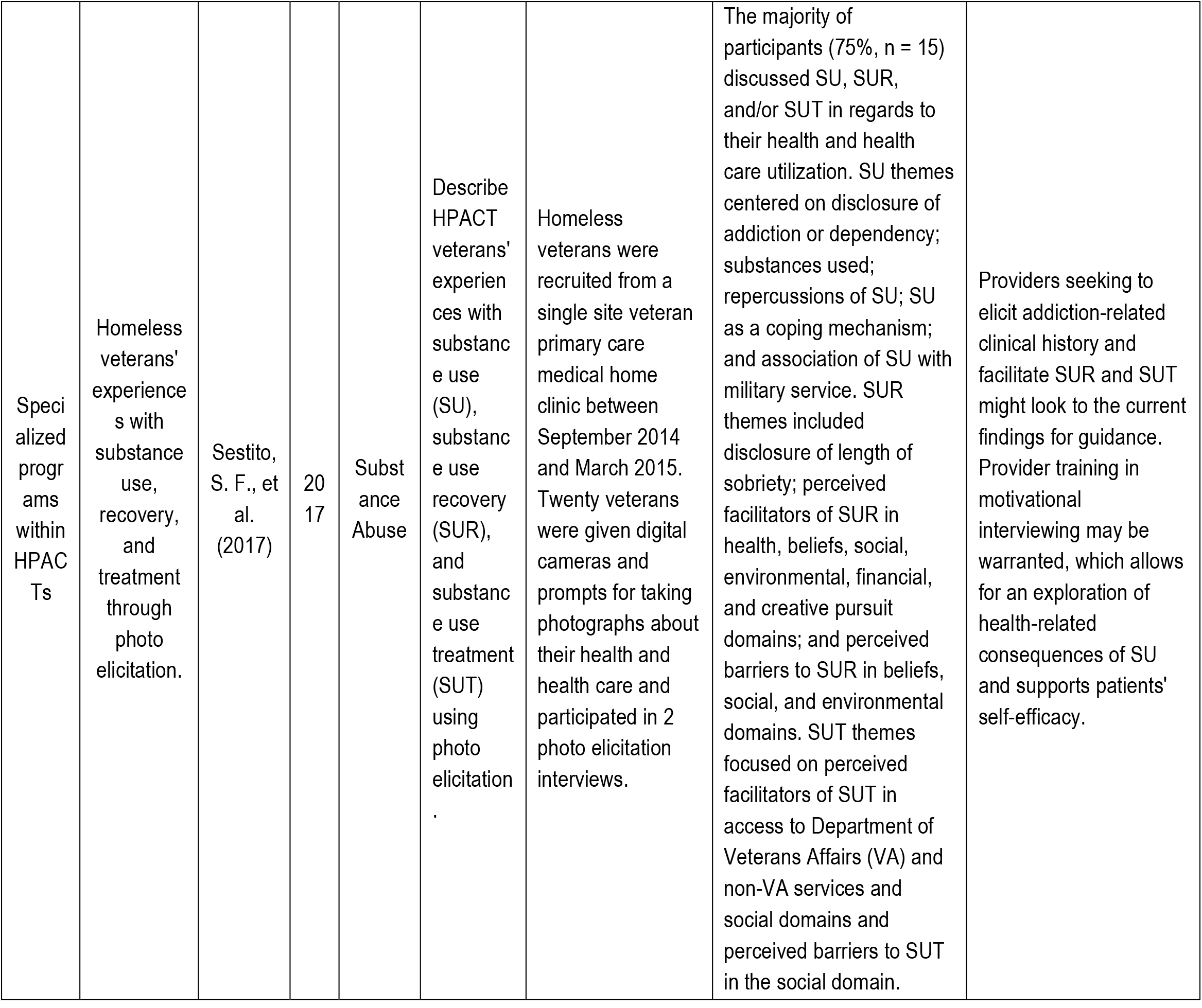
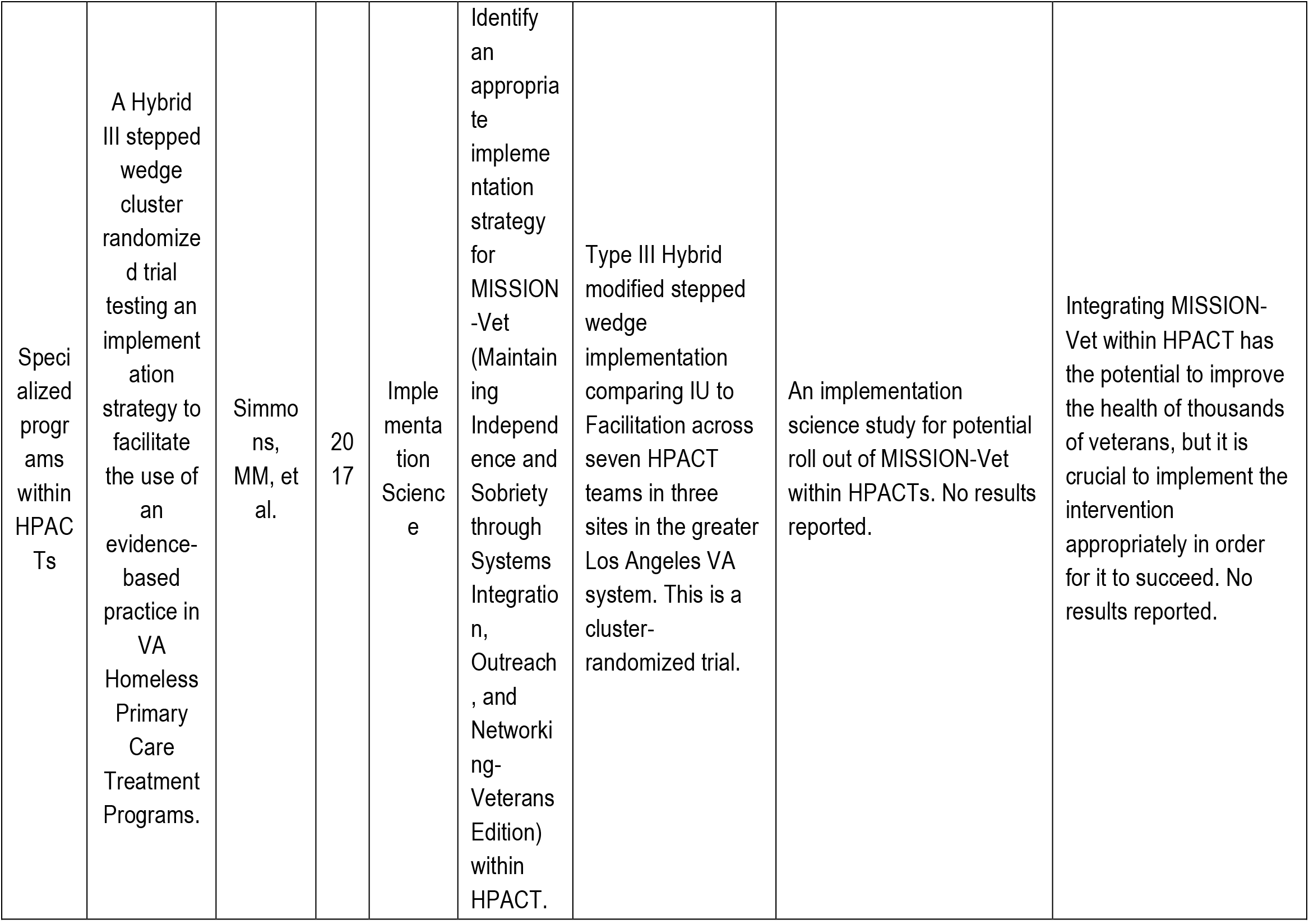
HPACT papers included in analysis

### (1) Early HPACT pilot implementations

Five papers were included that reported early implementation data. Four of these papers present original data from the pilot HPACT (known initially as Homeless-Oriented Primary Care Clinic) from the Providence VA Healthcare System. These Providence studies helped build the foundational evidence base for the creation and expansion of specialized PACT models, and specifically the HPACT model. In one study comparing the HPACT model to other specialized PACT models, O’Toole and colleagues used a quasi-experimental 6 month pre- and post-enrollment study tailoring patient-centered medical home interventions to four different high-risk groups and compared engagement and utilization – one of which was tailored to 71 homeless Veterans. This study demonstrated that a specific, homeless-oriented primary care model was feasible within the VA, and that pre-post comparison found a significant increase in primary care use although it was also accompanied by greater use of emergency department visits as well. (14) Furthermore, a larger randomized controlled trial (RCT), the only RCT to date about HPACTs with published data, by Providence VA researchers suggests that 185 out-of-treatment homeless Veterans could be effectively engaged in primary care, measured as any clinical visit in primary care during the study period, through a tailored outreach process through a Personal Health Assessment/Brief Intervention Arm (PHA/BI), a Clinic Orientation arm (HPACT), or both (PHA/BI+CO). (15) Both of these interventions significantly increased percentages of Veterans accessing primary care compared to a control arm of usual care, but had small numbers (around 40 in each intervention arm) of participants. O’Toole and colleagues in the Providence VA further observed that 79 Veterans experiencing homelessness had increased utilization of primary, mental health, substance abuse and ED care during the first 6 months of HPACT engagement as compared to 98 Veterans experiencing homelessness seen in traditional primacy care teams. (16) They found by tailoring primary care to homeless Veterans there were modest, statistically significant improvements in chronic disease management metrics (blood pressure, diabetes care, hyperlipidemia). (17) Some limitations of these original pilot studies are that they were small (in aggregate there were a total of about 441 non-unique Veterans experiencing homelessness in the four papers), at a single-site (Providence VA Healthcare System), and utilized different methodologies (1 quasi-experimental pre-post study, 1 retrospective cohort, 1 case-control nested cohort, and 1 prospective RCT). Despite these limitations, the data provided a foundational base for the national expansion of the HPACT model.

As the program moved from a single site to multiple sites, ability to assess fidelity to the original model became important. To better understand fidelity, O’Toole and colleagues in 2014 conducted an observational study of the first 33 VHA facilities with HPACTs with over 14,000 patients enrolled at the time. (12) Of the 33 sites studied, 82% provided hygiene care (on-site showers, hygiene kits, and laundry), 76% provided transportation, 55% had an on-site clothes pantry, and 42% had a food pantry and provided on-site meals or food assistance. Six-month patterns of acute-care use pre-enrollment and post-enrollment for 3,543 consecutively enrolled patients showed a 19.0% reduction in emergency department use and a 34.7% reduction in hospitalizations. Higher performance, as defined by the researchers as having greater reductions in ED and hospitalization use, was significantly associated with 1) higher staffing ratios than other sites, 2) integration of social supports and social services into clinical care, and 3) outreach to and integration with community agencies.

### (2) Association of HPACT Clinics with quality and utilization

A plurality of studies (nine) we included studied the association of HPACT clinics with quality of care and utilization of care by homeless Veterans. Four studies were surveys, three were retrospective observational studies, one a prospective quasi-experimental trial, and one a mixed-methodology qualitative interviews with descriptive data. None were prospective randomized control trials. Multiple studies focused on patient experience and engagement. Three studies suggest that care in an HPACT was associated with improved patient experience for homeless Veterans compared to traditional VA care settings (18, 19), especially for those Veterans with severe psychiatric symptoms. (20) Kertesz and colleagues in surveying about 600 homeless-experienced patients in 3 Veteran’s Affairs (VA) mainstream primary care settings, a Homeless-PACT VA clinic in California, and a highly tailored non-VA Health Care for the Homeless Program (HCHP) in Massachusetts reported that highly tailored primary care services were associated with decreased odds of unfavorable experiences in the domains of patient-clinician relationship, cooperation, and access or coordination compared to mainstream VA primary cares sites, whereas the VA HPACT site attained intermediate results. (21)

Between 2012 and 2014, O’Toole and colleagues studied 2 VHA medical centers to assess health services use, cost, and satisfaction over 12 months for a cohort of 266 homeless Veterans. (22) They found that compared with PACT patients, HPACT patients had more social work visits (4.6 vs 2.7 visits) and fewer emergency department (ED) visits for ambulatory care-sensitive conditions (0 vs 0.2 visits); a significantly smaller percentage of Veterans in HPACT were hospitalized (23.1% vs 35.4%) or had mental health–related ED visits (34.1% vs 47.6%). They also found significant differences in primary care provider–specific visits (HPACT, 5.1 vs PACT, 3.6 visits), mental health care visits (HPACT, 8.8 vs PACT, 13.4 visits), 30-day prescription drug fills (HPACT, 40.5 vs PACT, 58.8 fills), and use of group therapy (HPACT, 40.1% vs PACT, 53.7%). Annual costs per patient were significantly higher in the PACT group than the HPACT group ($37,415 vs $28,036), with higher primary care costs in the HPACT group but less cost from hospitalizations, based on managerial cost accounting estimates.

Regarding the effect of the HPACT model on utilization of care, in addition to the fidelity assessment performed by O’Toole and colleagues, several other studies have been performed. Some studies suggest that increased primary care engagement decreases ED and hospital utilization. Gundlapalli and colleagues showed that primary care treatment engagement can reduce ED visits, especially in the highest ED utilizers. (23) Jones and colleagues showed at a single HPACT site (Pittsburgh VAMC) that Veterans were more likely to have increased primary care visits and less likely to have ED visits following HPACT engagement. (24) Trivedi and colleagues observed that Veterans enrollment in HPACTs with significant dual use of Medicare and VA outpatient care was strongly associated with acute hospitalizations financed by Medicare and high fragmentation of care. (25) Other studies showed mixed results. Patel provided an early case study in 2013 at the VA greater Los Angeles Healthcare System of 47 Veterans who were assigned to the West LA HPACT for at least six months without any statistically significant change in acute ED utilization after six months despite significant increases in HPACT visits and SW encounters. (26) While limited by different methodologies and their observational nature, overall these studies seem to suggest that HPACTs increase engagement and utilization of primary care, and in the highest of utilizers, may be associated with a reduction in acute ED use and hospitalization.

### (3) Specialized programs within HPACTs

As the HPACT model has now been implemented in over 60 sites nationally, there have been innovative pilots and model variation described in the literature. Six studies were included in this category for analysis although all were of lower methodologic rigor. Several studies describe innovative staffing models. Pauly et al describe the integration of a mental health clinical pharmacy specialist in the South Texas HPACT that showed reduction in wait-time for psychiatric pharmacotherapy (27) whereas Yoon describes the use of Peer Mentors for Veterans experiencing Homelessness within HPACT and Non-HPACT clinics without significant impacts on Veteran utilization or cost. (28) Gabrielian and colleagues discuss the feasibility of a variation on the original HPACT model, describing a co-located HPACT clinic within the Emergency Department of the Greater Los Angeles VA Medical Center rather than in an ambulatory clinic. (29) Other innovations studied and described in the literature include implementation of food insecurity screening initiatives, (30) photo elicitation strategies for recovery, (31) and the MISSION-Vet program within an HPACT. (32) Each of these studies were included in the analysis because they contain original data; showing variation and ongoing innovation within the HPACT clinics.

## Discussion

The HPACT model has been implemented in VHA medical centers throughout the country and has great potential to engage Veterans experiencing homelessness in specialized access and services within primary care. Despite this rapid, widespread implementation, limited data exists about the program in the peer-reviewed literature. Our systematic review identified only 20 studies to date that provide an evidentiary base for evaluation of the program. The strongest evidence, obtained through the only randomized controlled trial of the HPACT model showed that it can increase patient engagement and increase primary care utilization. (14) In addition, there is modest observational data demonstrating an association between HPACT engagement and reduction in use of emergency care and acute hospitalization in certain high utilizing populations. Similarly, one observational study showed an association with HPACT program engagement and annual cost decreases from aforementioned decrease reduction in acute care. Importantly, HPACTs were not necessarily designed to improve housing outcomes per se so there is limited evidence in that respect, although several studies have shown that HPACTs can improve primary care engagement, which in theory could enhance health and housing outcomes. The impact of HPACTs on long-term housing instability has yet to be examined rigorously.

### Implication for Primary Care Settings and Health Care Policy

Overall, the HPACT model is a successful example of how an innovative new program can scale nationally within the VA. Within a few years from its single site pilot, over 60 sites nationally employ an HPACT model of care for Veterans experiencing homelessness. The rapid spread of the HPACT program does highlight the shift of framework from a “one-size-fits-all” primary care medical home model to a specialized model where segmentation and specialization of services may provide a slightly different model of care for vulnerable populations. Perhaps most striking from the majority of the studies, HPACTs are associated with an improved patient experience for Veterans experiencing homelessness compared to non-tailored primary care clinics. The implication for this is that better customizing and tailoring care to meet the specialized needs of this population is likely to result in better experience, higher engagement in treatment, and likely further retention in primary care. Additional research is clearly needed to better understand the effect of HPACT within VA, but also to better understand whether such a model could be implemented outside of a VA system, where other healthcare organizations now have an increased attention on improving social determinants of health.

### Limitations and Strengths

This study has significant limitations. First, there may have been bias in our search and study selection strategy. Our search was limited to peer-reviewed publications and therefore excluded non-peer reviewed literature about HPACTs and internal, non-published quality-improvement data about HPACTs that exist such as within the National VA VISN Support Services Center. We attempted to mitigate this bias by conducting our search according to Preferred Reporting Items for Systematic Reviews and Meta-Analyses (PRISMA) guidelines and standard accepted databases, but it is likely that there exists more evidence about the HPACT program outside of the published literature. Furthermore, regarding potential selection bias, the two reviewers of studies both work clinically in HPACT clinics and may have implicitly excluded or interpreted studies towards a positive associations. Ten studies were excluded from analysis because they did not evaluate the HPACT program but evaluated them in combination with other VHA homeless programs such as HUDVASH and supportive housing interventions. It is possible that these exclusions removed important data from the review; however, we limited the scope of this review to interventions specific to HPACTs.

Second, one of the largest challenges with conducting this study was the issue of fidelity to the HPACT model. In almost all of the published studies, is not clear and consistent to the authors what the HPACT team is – i.e. whether there are consistent staffing ratios, consistent training between sites, and how geographic factors may play an outsized role in accessibility to services. Perhaps most importantly, its not clear that there are predetermined standard performance metrics and outcomes that the HPACTs are seeking to achieve and whether HPACTs consistently use data reports to manage towards these specific outcomes. Due to the significant internal variability of these programs, generalizability of the HPACT model is significantly impaired. Regarding costs, studies suggesting cost benefit of HPACT do not clarify whether they include HPACT staffing costs, and should be noted that the data used for non-VA use and cost are only estimates based on Fee Basis files.

Lastly, the methodology and quality of the studies makes comparison of studies challenging. In particular, numerous observational trials were subject to multiple confounders, in particular, the significant growth of homeless services in VA at the same time as the implementation of the HPACT programs, that may increase patient engagement to VA services independently of the HPACT program.

## Conclusion

People experiencing homelessness are high utilizers of high cost services in our health care system and unfortunately also have high levels of morbidity compared to the non-homeless population. The VHA should be applauded for implementing a specialized primary care model, the HPACT, to address these issues for veterans experiencing homelessness. Our review shows that the HPACT model has been successfully implemented in VHA medical centers throughout the country with multiple studies showing increased primary care engagement and improved patient experience. However, to date, published studies evaluating the HPACT model are limited and further studies are needed to refine our understanding of the impact that HPACTs have on quality, utilization and whether the model can be implemented outside the VHA system.

## Supporting information

Supplemental Search Strategy and PRISMA

## Data Availability

Systematic review performed from literature search in appendix.

## References

(1) Hwang SW, Orav J, O’Connell JJ, et al. Causes of death in homeless adults in Boston. Ann Intern Med. 1997;126(8):625–628.

(2) Baggett TP, Hwang SW, O’Connell JJ, et al. Mortality among homeless adults in Boston: shifts in causes of death over a 15-year period. JAMA Intern Med. 2013; 173(3):189–195.

(3) Kushel MB, Perry S, Bangsberg D, et al. Emergency department use among the homeless and marginally housed: results from a community-based study. Am J Public Health. 2002;92(5):778–784

(4) Kushel MB, Vittinghoff E, Haas JS. Factors associated with the health care utilization of homeless persons. JAMA. 2001;285(2):200–6

(5) Institute of Medicine/Committee on Health Care for Homeless People. Homelessness, Health, and Human Needs. Washington, DC: National Academy Press; 1988

(6) O’Toole TP, Gibbon JL, Hanusa BH, et al. Utilization of health care services among subgroups of urban homeless. J Health Polit Policy Law. 1999;24(1): 91–114

(7) Donaldson MS, Vanselow NA, KD Yordy, editors. Institute of Medicine Committee on the Future of Primary Care. Defining primary care: an interim report. Washington, DC: National Academies Press; 1994

(8) O’Toole TP, Johnson EE, Redihan S, et al. Needing primary care but not getting it: the role of trust, stigma and organizational obstacles reported by homeless Veterans. J Health Care Poor Underserved. 2015;26(3):1019–1031.

(9) US Department of Veterans Affairs, Office of Public and Intergovernmental Affairs. Secretary Shinseki details plan to end homelessness for Veterans. http://www.va.gov/opa/pressrel/pressrelease.cfm?id=1807 Accessed 1/7/2020

(10) O’Toole TP, Buckel L, Bourgault C, et al. Applying the chronic care model to homeless Veterans: effect of a population approach to primary care on utilization and clinical outcomes. American journal of public health 2010;100:2493–9.

(11) O’Toole TP, Johnson EE, Aiello R, et al. Tailoring Care to Vulnerable Populations by Incorporating Social Determinants of Health: the Veterans Health Administration’s “Homeless Patient Aligned Care Team” Program. Prev Chronic Dis 2015;30(7):886–98.

(12) O’Toole TP, Johnson EE, Aiello R, et al. Tailoring care to vulnerable populations by incorporating social determinants of health: the Veterans health administration’s “Homeless Patient Aligned Care Team” program. Prev Chronic Dis. 2016;13:150567.

(13) Moher D, Liberati A, Tetzlaff J, et al. Preferred reporting items for systematic reviews and meta-analyses: the PRISMA statement. BMJ 2009; 339: b2535.; Annals of Internal Medicine. 151. 264. 10.7326/0003-4819-151-4-200908180-00135.

(14) O’Toole TP, Pirraglia PA, Dosa D, et al. Building care systems to improve access for high-risk and vulnerable Veteran populations. J Gen Intern Med. 2011;26 Suppl 2(Suppl 2):683–688. doi:10.1007/s11606-011-1818-2

(15) O’Toole TP, Johnson EE, Borgia ML, et al. Tailoring outreach efforts to increase primary care use among homeless Veterans: results of a randomized controlled trial. J Gen Intern Med. 2015;30:886–898.

(16) O’Toole TP, Bourgault C, Johnson EE, et al. New to care: demands on a health system when homeless Veterans are enrolled in a medical home model. Am J Public Health. 2013;103 Suppl 2(Suppl 2):S374–S379. doi:10.2105/AJPH.2013.301632

(17) O’Toole TP, Buckel L, Bougault C, et al. Applying the chronic care model to homeless Veterans: effect of a population approach to primary care on utilization and clinical outcomes. Am J Public Health. 2010;100(12):2493–9.

(18) Jones AL, et al. Providing positive primary care experiences for homeless Veterans through tailored medical homes: the Veterans health administration’s homeless patient aligned care teams. Med Care. 2019;57(4):270–8.

(19) Jones, AL, Hausmann, LRM, Kertesz, SG, et al. (2018). Differences in experiences with care between homeless and nonhomeless patients in Veterans affairs facilities with tailored and nontailored primary care teams. Medical Care, 56, 610–618.

(20) Chrystal, JG., Glover, DL, Young, AS, et al. (2015). Experience of primary care among homeless individuals with mental health conditions. PLoS One, 10(2), e0117395

(21) Kertesz SG, Holt CL, Steward JL, et al. Comparing homeless persons’ care experiences in tailored versus nontailored primary care programs. Am J Public Health, 103 (suppl 2) (2013), pp. S331–S339

(22) O’Toole TP, Johnson EE, Borgia M, et al. Population-tailored care for homeless Veterans and acute care use, cost, and satisfaction: A prospective quasi-experimental trial. Preventing chronic disease. 2018;15:E23.

(23) Gundlapalli AV, Redd A, Bolton D, et al. Patient-aligned care team engagement to connect Veterans experiencing homelessness with appropriate health care. Med Care. 2017;55:S104–10.

(24) Jones AL, Thomas R, Hedayati DO, et al. Patient predictors and utilization of health services within a medical home for homeless persons. Substance abuse. 2018;39(3):354-360. PMID: 29412071.

(25) Trivedi AN, Jiang, L, Johnson EE, et al. Dual Use and Hospital Admissions Among Veterans Enrolled in the VA’s Homeless Patient Aligned Care Team. Health Services Research 2018; 53 (6 Pt 2): 5219–37.

(26) Patel, BI, Complex Care Management to Decrease Emergency Department Utilization: A Case Study of the Homeless Patient Aligned Care Team Demonstration Project at VA Greater Los Angeles Healthcare—Escholarship. UCLA, 2013. Available online: https://escholarship.org/uc/item/1681c184

(27) Pauly JB, Moore TA, Shishko I. Integrating a mental health clinical pharmacy specialist into the Homeless Patient Aligned Care Teams. Ment Health Clin [Internet]. 2018; 8 (4): 169–74.

(28) Yoon J, Lo J, Gehlert E, et al.: Homeless Veterans’ use of peer mentors and effects on costs and utilization in VA clinics. Psychiatric Services 2017; 68:628–631.

(29) Gabrielian S., Chen JC, Minhaj BP, et al. Feasibility and acceptability of a co-located homeless-tailored primary care clinic and emergency department, J Prim Care Commun Health, 8 (4) (2017), pp. 338–344

(30) O’Toole TP, Roberts CB, Johnson EE. Screening for food insecurity in six Veterans administration clinics for the homeless, June-December 2015. Prev Chronic Dis. 2017;14:4.

(31) Sestito SF, Rodriguez KL, Saba SK, et al. Homeless Veterans’ experiences with substance use, recovery, and treatment through photo elicitation. Subst Abus. 2017;38(4):422–31.

(32) Simmons MM, Gabrielian S, Byrne T, et al. A Hybrid III stepped wedge cluster randomized trial testing an implementation strategy to facilitate the use of an evidence-based practice in VA Homeless Primary Care Treatment Programs. Implement Sci. 2017;12:46.

