## Supplemental Search Strategy and PRISMA for "Primary Care for Homeless Veterans: A Systematic Review of the Homeless Patient Aligned Care Team (HPACT)"

Table S-1. Search Strategy

**Ovid MEDLINE(R) Epub Ahead of Print, In-Process & Other Non-Indexed Citations, Ovid MEDLINE(R) Daily and Ovid MEDLINE(R) <1946 to Present>**

| Search history sorted by search number ascending | | | |
| --- | --- | --- | --- |
| **#** | **Subject Term(s)** | **Results** | **Type** |
| 1 | HPACT.mp. | 7 | Advanced |
| 2 | Homeless patient aligned care team.mp. | 7 | Advanced |
| 3 | 1 or 2 | 9 | Advanced |
| 4 | United states department of Veterans affairs.mp. OR exp “united states department of Veterans Affairs” | 7812 | Advanced |
| 5 | Exp Veterans health OR exp hospitals, Veterans OR Veterans | 21664 | Advanced |
| 6 | 4 OR 5 | 25313 | Advanced |
| 7 | Exp homeless persons | 8461 | Advanced |
| 8 | 6 and 7 | 424 | Advanced |
| 9 | exp Ambulatory care OR exp delivery of health care OR exp patient-centered care OR exp primary health care OR exp health services needs and demand | 1161066 | Advanced |
| 10 | exp Health Services Accessibility | 106277 | Advanced |
| 11 | exp Delivery of health Care, Integrated | 12208 | Advanced |
| 12 | 9 OR 10 OR 11 | 1161066 | Advanced |
| 13 | 8 AND 12 | 156 | Advanced |

**Ovid EMBASE 1974 to present**

| Search history sorted by search number ascending | | | |
| --- | --- | --- | --- |
| **#** | **Subject Term(s)** | **Results** | **Type** |
| 1 | HPACT.mp. | 8 | Advanced |
| 2 | Homeless patient aligned care team.mp. | 11 | Advanced |
| 3 | 1 OR 2 | 13 | Advanced |
| 4 | United states department of Veterans affairs.mp. | 153 | Advanced |
| 5 | exp Veterans health | 3089 | Advanced |
| 6 | Hospitals Veterans | 28 | Advanced |
| 7 | Veterans hospitals.mp. | 76 | Advanced |
| 8 | 6 OR 7 | 104 | Advanced |
| 9 | Veterans.mp. OR exp Veteran | 45038 | Advanced |
| 10 | 4 OR 5 OR 6 OR 7 OR 8 OR 9 | 45038 | Advanced |
| 11 | sxp homeless person | 1655 | Advanced |
| 12 | 10 and 11 | 105 | Advanced |

**PsycINFO 1967 to present**

| Search history sorted by search number ascending | | |  |
| --- | --- | --- | --- |
| # | **Subject Term(s)** | **Results** | **Type** |
| 1 | HPACT.mp. | 1 | Advanced |
| 2 | Homeless patient aligned care team.mp. | 3 | Advanced |
| 3 | 1 OR 2 | 3 | Advanced |
| 4 | exp military Veterans OR united states department of Veterans affairs.mp. | 13036 | Advanced |
| 5 | exp health care utilization OR exp health care services OR exp health care delivery OR exp primary health care OR Veterans health.mp. | 199538 | Advanced |
| 6 | 4 OR 5 | 209626 | Advanced |
| 7 | Homeless Veterans.mp. | 271 | Advanced |
| 8 | 6 AND 7 | 236 | Advanced |
| 9 | limit 8 to (all journals and English language) | 194 | Advanced |
| 10 | 3 OR 9 | 196 | Advanced |

Figure S-1. PRISMA flow diagram

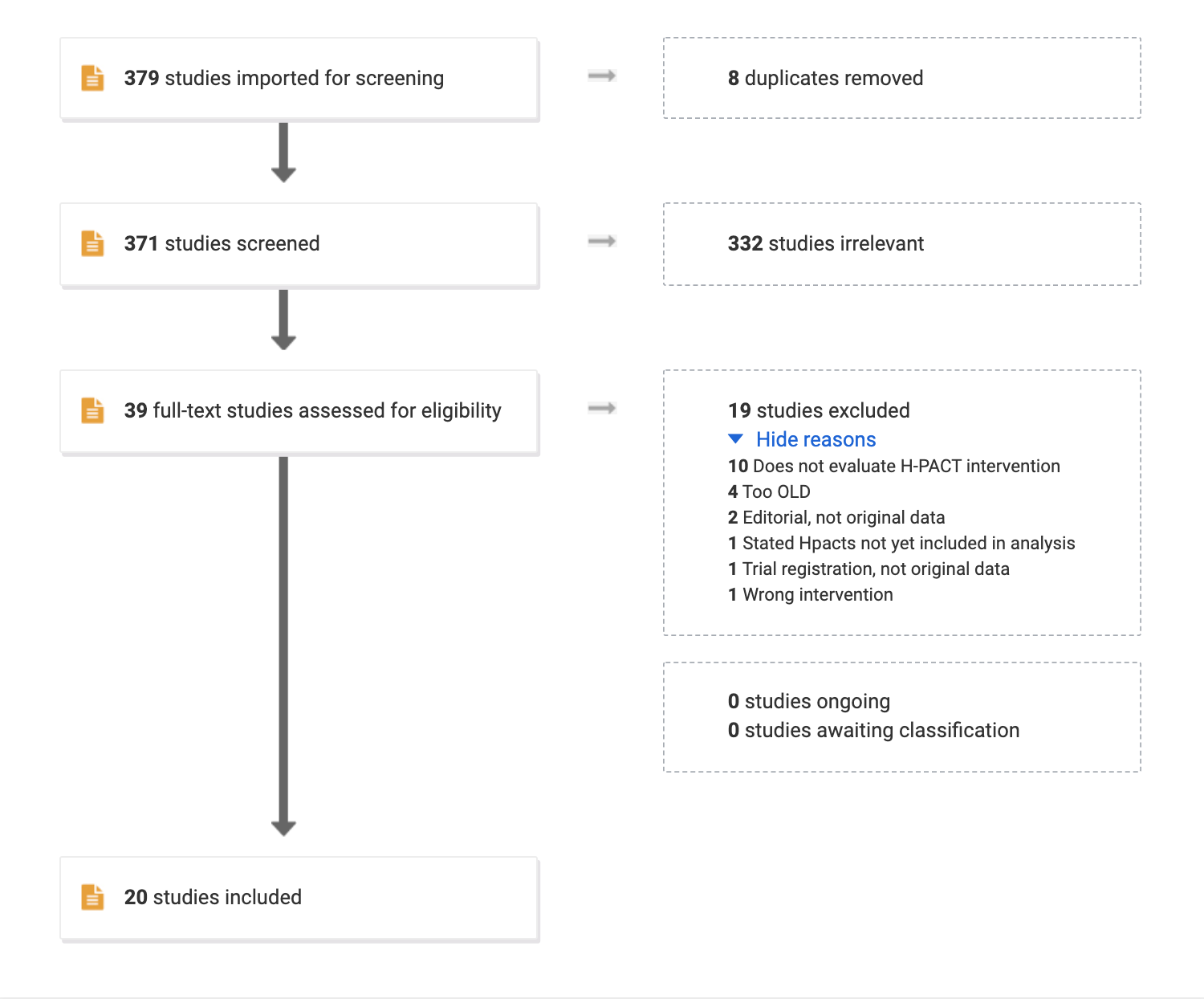
